# Angle-angle diagrams in the assessment of locomotion in persons with multiple sclerosis: A preliminary study

**DOI:** 10.1101/2022.01.09.22268813

**Authors:** Riccardo Di Giminiani, Davide Di Lorenzo, Luca Russo, Stefano La Greca, Francesco Masedu, Rocco Totaro, Elvira Padua

**Affiliations:** Department of Biotechnological and Applied Clinical Sciences-University of L’Aquila, Italy; Italian University Line-IUL, Firenze, Italy; Centre for the Diagnosis and Treatment of Demyelinating Diseases, Hospital of L’Aquila-Sulmona-Avezzano, Italy; Department of Human Science and Promotion of Quality of Life-San Raffaele Open University Rome

**Keywords:** Gait analysis, walking, knee-angle/hip-angle relationships, EMG activity, neurodegenerative

## Abstract

Gait analysis is clinically relevant in persons with multiple sclerosis (PwMS) and consists of several joint angular displacement-time relationships and spatiotemporal parameters. However, it lacks representation by means of diagrams in which knee angle-hip angle and knee angle-ankle angle variations are plotted against each other at the same instants of time. Three-dimensional kinematic analysis was performed on 20 subjects (10 PwMS/10 healthy controls, HCs), and the knee-angle/hip-angle and knee-angle/ankle-angle diagrams of both lower limbs were determined in the sagittal plane while walking on a motorized treadmill. The area (a quantifier of conjoint range of motion) and the perimeter (a quantifier of coordination) of angle-angle diagram loops were calculated. PwMS showed reduced knee-angle/ankle-angle loops compared to HCs (P<0.05, ES = 0.80), whereas the hip-angle/ankle-angle loops between the PwMS and HCs was not significant (P> 0.05). Similarly, the activation of leg muscles showed significant differences between PwMS and HCs (p ranged from 0.05-to 0.001; ES ranged from 1.30 to 1.89). The results indicate that the proposed knee-angle/hip-angle diagram is feasible and could be applied as a reliable tool in future studies aimed at assessing the acute and long-term effects of specific exercise programmes and/or pharmacological treatment in PwMS.

## Introduction

Multiple sclerosis (MS) is a chronic disease of the nervous system characterized by demyelination of the axonal myelin sheath that can occur in different areas, with consequent axonal damage. This means that the symptomatology is variable and progressive (1–3). Early patients affected by MS present with gait disorders associated with muscular weakness and spasticity, resulting in a decrease in quality of life, which can be monitored by the expanded disability status scale (EDSS). The evaluation of a patient’s quality of life is one of the most effective methods in monitoring the clinical course of the disease (4); as the EDSS reports, gait impairments and balance disorders are two of the major aspects from the fourth level up (5).

Gait analysis in people affected by multiple sclerosis reveals some shared traits, such as lower walking speed, frequency and step length than healthy people and higher double support duration, step width and stride time than healthy people (6). Moreover, it has already been shown that in the MS population, a decrease in the range of motion (ROM) occurs in hip, knee and ankle joints (7). However, the data obtained through gait analysis usually consist of relationships between joint angular variations and time, which gives us information that is not easy to interpret in clinical assessments (6–8). Conversely, angle-angle diagrams plot the angles at adjacent joints against each other rather than plotting angles versus time. For example, in the sagittal plane of motion, the hip-angle/knee-angle diagram displays the joint range of hip flexion-hyperextension along the X-axis and the range of knee flexion along the Y-axis. Therefore, the area delimited in this diagram depends on hip and knee angles, which correspond to the total conjoint range of angular motion performed by the two joints during one complete gait cycle (9).

When changes occur in both joints, a corresponding perimeter variation is determined in the angle-angle diagram. If any joint rotation is uncoordinated, the perimeter will increase, while the conjoint range of motion (area) may remain approximately constant. Although perimeter is a function of both angles, when the variation involves one joint more than the other, a change in the perimeter may define a pattern describing a robot-like gait in which only one joint rotates and the other joint is fixed. In other words, the perimeter may reflect the coordination between two joints during gait (9). Therefore, it might also be interesting to analyse gait from a quantitative perspective through three joints (hip, knee and ankle) that define two angle-angle diagrams in the lower limb (9,10), which is an approach that is still not applied for gait analysis in persons with multiple sclerosis (PwMS). The hip-angle/knee-angle and the knee-angle/ankle-angle loops can provide quantitative information such as co-joint range of motion and coordination by calculating the areas and perimeters, respectively. It is also possible to obtain qualitative information observing the “shape” of the diagrams.

We hypothesized that the main characteristics of the hip-angle/knee-angle loops and the knee-angle/ankle-angle loops (i.e., area and perimeter) may represent a new approach to observing gait pattern alterations in people affected by multiple sclerosis, providing additional information about the status/progress of the disease and for monitoring pharmacological treatment and/or kinesiological intervention. In addition, gait measures can be synchronized with surface electromyography (sEMG) of the lower limb muscles to describe the activation in proximal and distal leg muscles during walking (11).

The first aim of this study was to determine the areas and perimeters of the hip-angle/knee-angle diagrams and knee-angle/ankle angle diagrams. The diagrams were obtained in a sagittal plane of motion in PwMS and healthy controls (HCs) while walking at a defined speed. Second, we examined the usefulness of angle-angle diagrams in relation to the sEMG activity of the extensor and flexor muscles of the lower extremities.

## Materials and Methods

### Participants

PwMS with a low level of disability that allowed them to walk without any external support were recruited from the Centre for the Diagnosis and Treatment of Demyelinating Diseases in the Hospital of L’Aquila-Sulmona-Avezzano (8 females/2 males: age 52.7±10.3 years; stature 165.0±8.8 cm; body mass 67.2±11.2 kg; body mass index 24.8±4.5 kg· m^-2^). HCs who had similar characteristics to those of the PwMS were recruited (8 females/2 males: age 52.3±10.9 years; stature 165.5±7.3 cm; body mass 60.7±9.5 kg; body mass index 22.11±2.7 kg· m^-2^). The inclusion criterion for PwMS was a multiple sclerosis diagnosis with an EDSS score of 4/5, while the exclusion criteria were other diseases of the neuromuscular system. For the HCs, the inclusion criteria were an age of between 50 and 70 years and motor skills that did not prejudice walking, while the exclusion criteria were growth and development anomalies, pharmacological therapy and present or past history of musculoskeletal and nervous diseases. The local ethics committee named Internal Review Board of the University of L’Aquila, Abruzzo, Italy (Protocol number 13/2021; www.univaq.it/en/section.php?id=1527) approved the study. Following all the participants gave their written informed consent according to the Helsinki declaration.

### Surface EMG

sEMG was recorded using bipolar electrodes (Ambu Neuroline 720 00-S/25, electrode diameter 45×22, interelectrode distance 2,5 cm, Ambu A/S, Baltorpbakken, Denmark) in four muscles: vastus lateralis (VL), bicep femoris (BF), tibialis anterior (TA) and lateral gastrocnemius (LG). Before placing the electrodes on the muscle belly, the skin was shaved and then cleaned with an alcohol solution to minimize impedance (<5 kΩ) following the SENIAM recommendation for surface electromyography (12). The electrodes and modules were fixed on the thigh and shank with an elastic band to prevent motion artefacts. The wireless data synchronization unit (MuscleLab 6000, Ergotest-innovation, Porsgrunn, Norway) characteristics were as follows: a built-in radio frequency (RF) module ML6RFM02 with 2.4 GHz and 1 mW, and a typical wireless range in open space of 20 m. The modules of the MuscleLab characteristics were as follows: RF characteristics of 2.4 GHz, 1 mW; sample rate of 1 kHz; high-pass filter of 20 Hz; bandwidth of 20 Hz-500 Hz; input signal range of ±5 mVp-p; noise of 14 µVp-p (2.2 µVrms); and resolution of 0.33 µV/bit. The RMS was determined in ten successive stance phases for the dominant leg for the HCs (1 and 3 km/h) and for the weakest leg in the PwMS (1 km/h). Ten stance phases were selected in the middle part of the kinematic walk recording.

### Gait analysis

The participants walked on a motorized treadmill, and kinematic analysis was performed using four optoelectronic cameras positioned around the tapis and connected to the SMART Motion Capture System (BTS, Bioengineering, Milano). Reflecting markers were positioned in correspondence with the greater trochanter, the lateral femoral condyle, lateral malleolus and fifth metatarsus (Fig 1A). The walking speed was determined through the timed 25-foot walk (T25FW)(13), which consisted of measuring the required time to walk a distance of 25 feet (8 metres) as quickly as possible without any external support. On the treadmill, the experimental group walked at a 1-km/h speed for a duration of 30 seconds, while the HCs performed for the same duration at 1 and 3 km/h.

**Fig 1.**
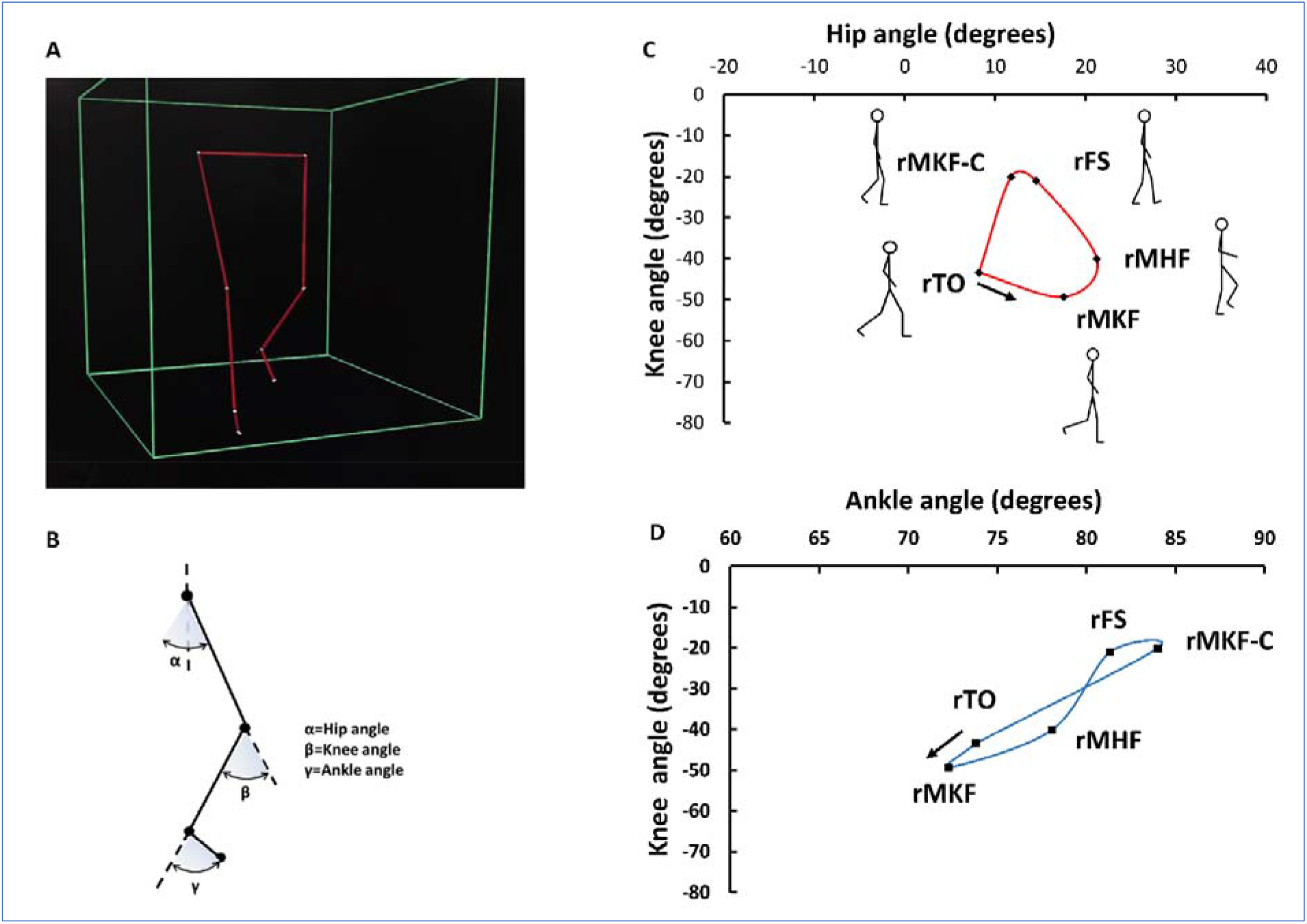
Determination of angle-angle diagram of a person with MS. (A) Lower limb reconstruction obtained by SMART Motion Capture System (BTS, Bioengineering, Milano). (B) Stick figure of the lower limb with hip angle (α), knee angle (β) and ankle angle (γ). (C) Angle-angle diagram of the right leg of a person with MS (rTO= right take off, rMKF= maximum knee flexion, rMHF= maximum gip flexion, rFS= right foot strike, rMKF-C= right maximum knee flexion during the contact phase).

The obtained data were analysed using the SMART Analyser (BTS, Bioengineering, Milano). Three angles and their trends over time were plotted in the sagittal plane motion, namely, the knee angle, ankle angle (relative) and hip angle (absolute). The relative angles are the angle formed by the extension of the thigh axis and shank axis (knee angle) and the angle formed by the shank axis and foot, while the absolute hip angle is formed by the thigh axis and the vertical hip passing through the hip (Fig 1B).

Two gait cycles of each leg were used to analyse the reliability (test-retest), and then the average values were calculated to plot two angle-angle relations, i.e., the weakest and strongest leg for PwMS and the dominant and nondominant leg for HC. The selected cycles were as central in the test duration as possible so that the participants could adapt themselves to the speed of the treadmill. The angle-angle diagrams were plotted according to the study provided by Cavanagh (14), in which five points to interpole were chosen: toe off (TO), maximum knee flexion (MKF), maximum hip flexion (MHF), foot strike (FS), maximum knee flexion during the contact phase (MKF-C); furthermore, concerning MKF, the frame with the minimum value of the knee angle was selected, while for the MHF, the frame with the minimum value of the hip angle was selected, and for the MKF-C, the frame with the minimum value of the knee angle during the contact phase was selected (Figs 1C, 1D). For the TO and the FS, the considered frames corresponded to those in which the value of the X vector of velocity changed from negative to positive and vice versa (15). The areas and perimeters of the angle-angle diagrams were calculated by using AutoCAD 2017 (Autodesk, San Rafael, California, USA), and the dimensions of the diagrams were the same for all the participants (PwMS and HCs).

### Statistical analysis

Areas and perimeters of the angle-angle diagrams and EMG activity comparisons were performed with the Wilcoxon signed-rank test/two-tailed test in paired samples and with the Mann–Whitney test/two-tailed test in unpaired samples. Statistical significance was set at α = 0.05, and the meaningfulness of significant outcomes was estimated by calculating the ES of Cohen. The intrasession reliability of the EMG and angle data was quantified using the intraclass correlation coefficient (ICC) (16). ICC values less than 0.50 are defined as “poor,” those from 0.50 to 0.69 are defined as “moderate,” those from 0.70 to 0.89 are defined as “high,” and those greater than 0.90 are defined as “excellent” (17). The variability of areas and perimeters between and within the subjects was estimated by calculating the coefficient of variation (CV, %) in each leg. The analysis was executed using the statistical software XLSTAT 2013.2.07 (Addinsoft; New York, USA).

## Results

### Reliability of the Measurements

The intraclass coefficient correlation (ICC) of the hip, knee and ankle angles in two consecutive gait cycles was equal to 0.86 in PwMS (the confidence interval 95%; the lower confidence limit was equal to 0.67; the upper confidence limit was equal to 0.95) and 0.98 in HCs (the confidence interval was 95%; the lower confidence limit was equal to 0.95; the upper confidence limit was equal to 0.99). In PwMS, the ICCs of the EMG_RMS_ values recorded in the repeated contact phases were 0.94, 0.94, 0.94, 0.76 and 0.95 for VL, BF, TA and LG, respectively. The ICC values in the HCs were 0.97-0.85 (VL 1-3 km/h), 0.92-0.91 (BF 1-3 km/h), 0.83-0.75 (TA 1-3 km/h) and 0.75 (LG 1-3 km/h).

### Angle-Angle Diagrams

The area of the knee-angle/ankle-angle loops between PwMS vs. HC-1 at the same speed showed significant differences (P<0.05, ES = 0.80, Fig 2A), whereas the hip-angle/ankle-angle loops did not show significant differences (P> 0.05). The perimeter differences at a speed of 1 km/h tended to be significant between the knee-angle/ankle-angle loops (P = 0.086, ES = 0.53) but not between the hip-angle/knee-angle loops (P = 0.678) (Fig 2B). The area and perimeter comparisons within the HCs when they walked at different speeds (HC-1 vs. HC-3) were significant either between the hip-angle/knee-angle loops (P<0.001, ES ranged from 2.0 to 3.1) or between the knee-angle/ankle-angle loops (P< 0.001, ES ranged from 0.93 to 1.70) (Figs 2A, 2B). The variability of two consecutive gait cycles (CV, %) between and within the subjects is reported in Table 1 and Figs 3-5.

**Table 1.**
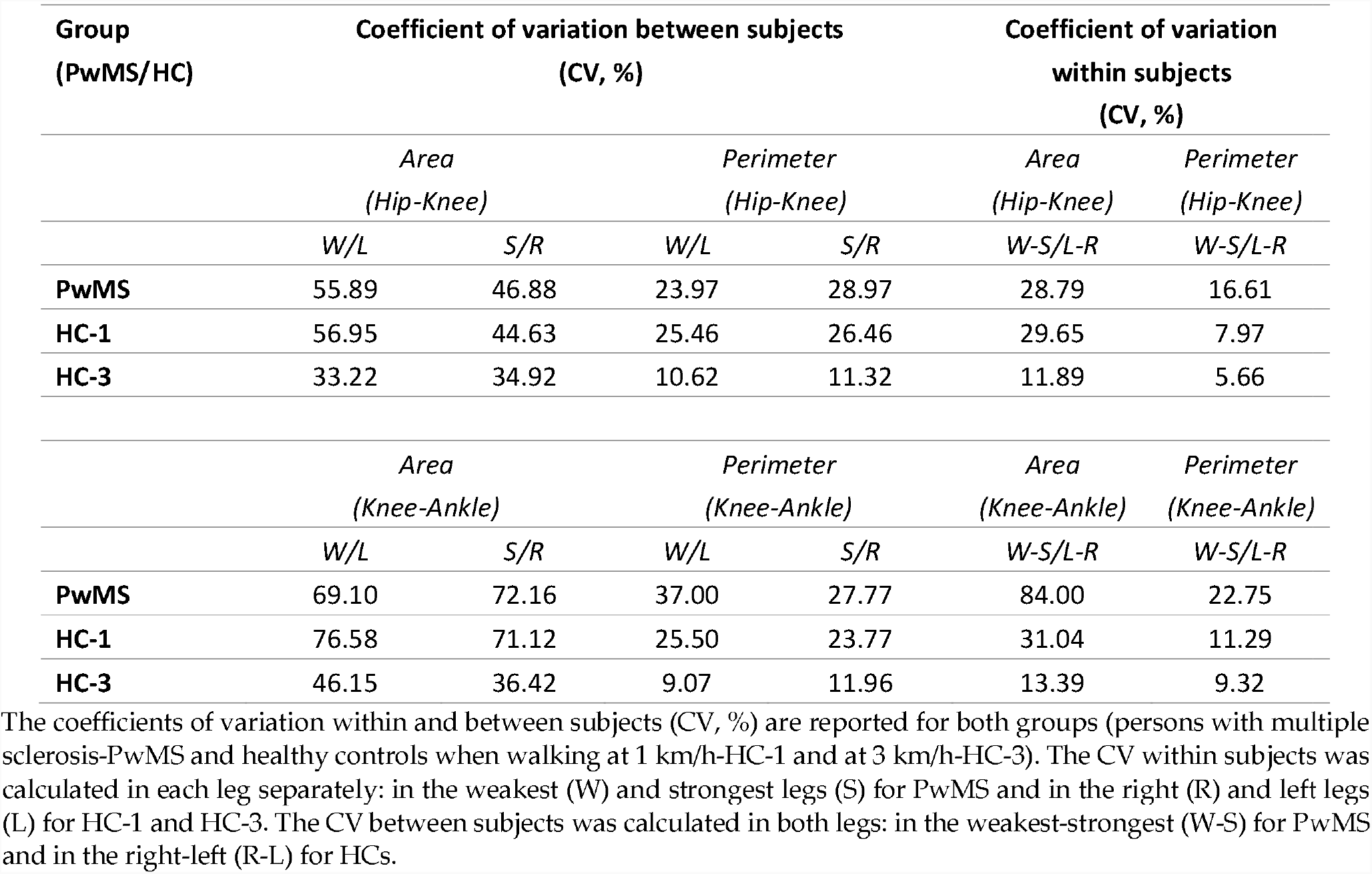
Coefficients of variation.

**Fig 2.**
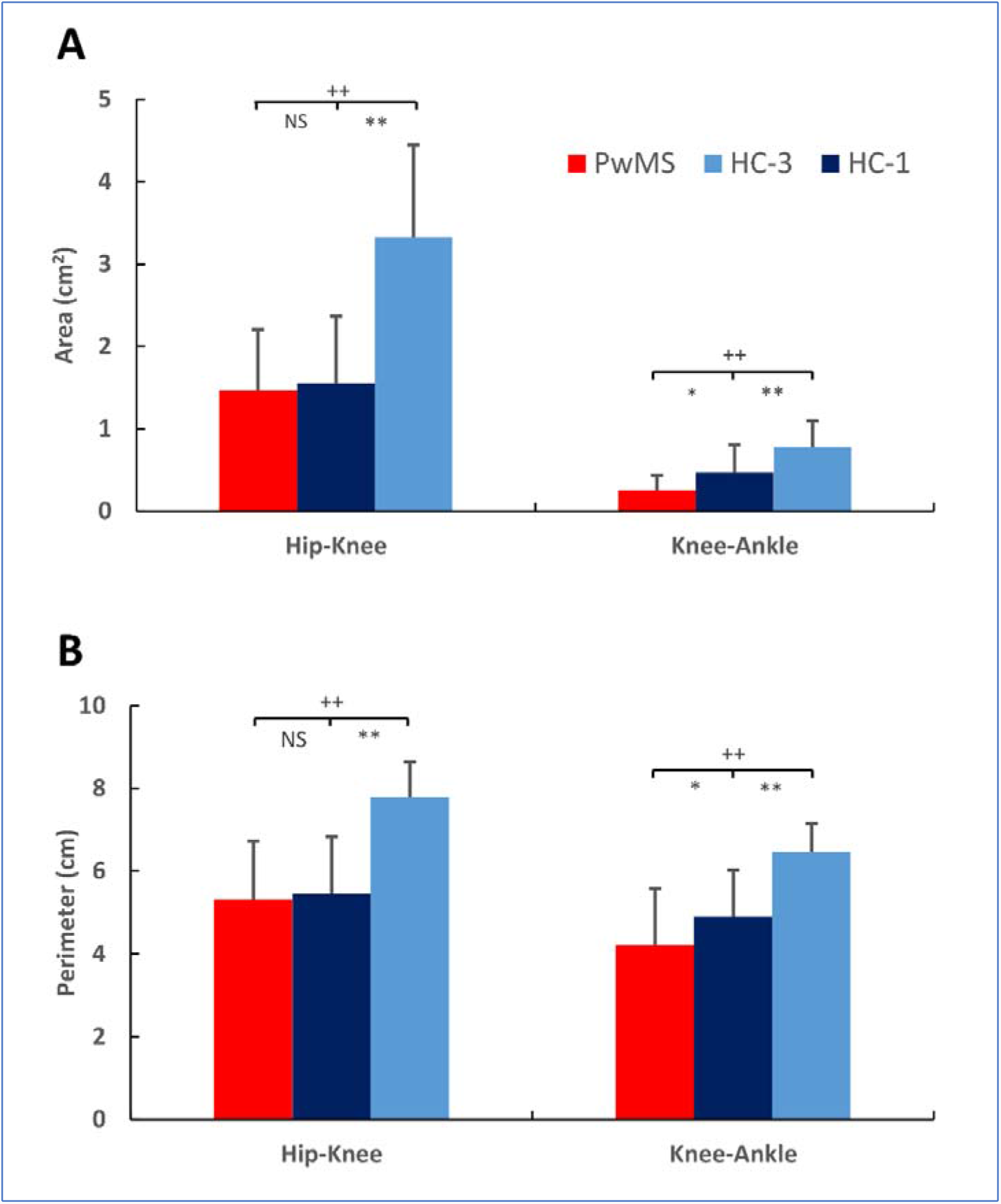
Values (SD) of areas (A) and perimeters (B) in the hip-knee and knee-ankle diagrams are reported in both groups (PwMS, HC-1=1 km/h and HC-3=3 km/h). *Difference between PwMS and HC-1 (p<0.05); ^++^Difference between HC-3 and PwMS (p<0.001); ^**^Difference between HC-3 and HC-1 (p<0.001); NS = no significant difference between HC-1 and PwMS (p>0.05).

**Fig 3.**
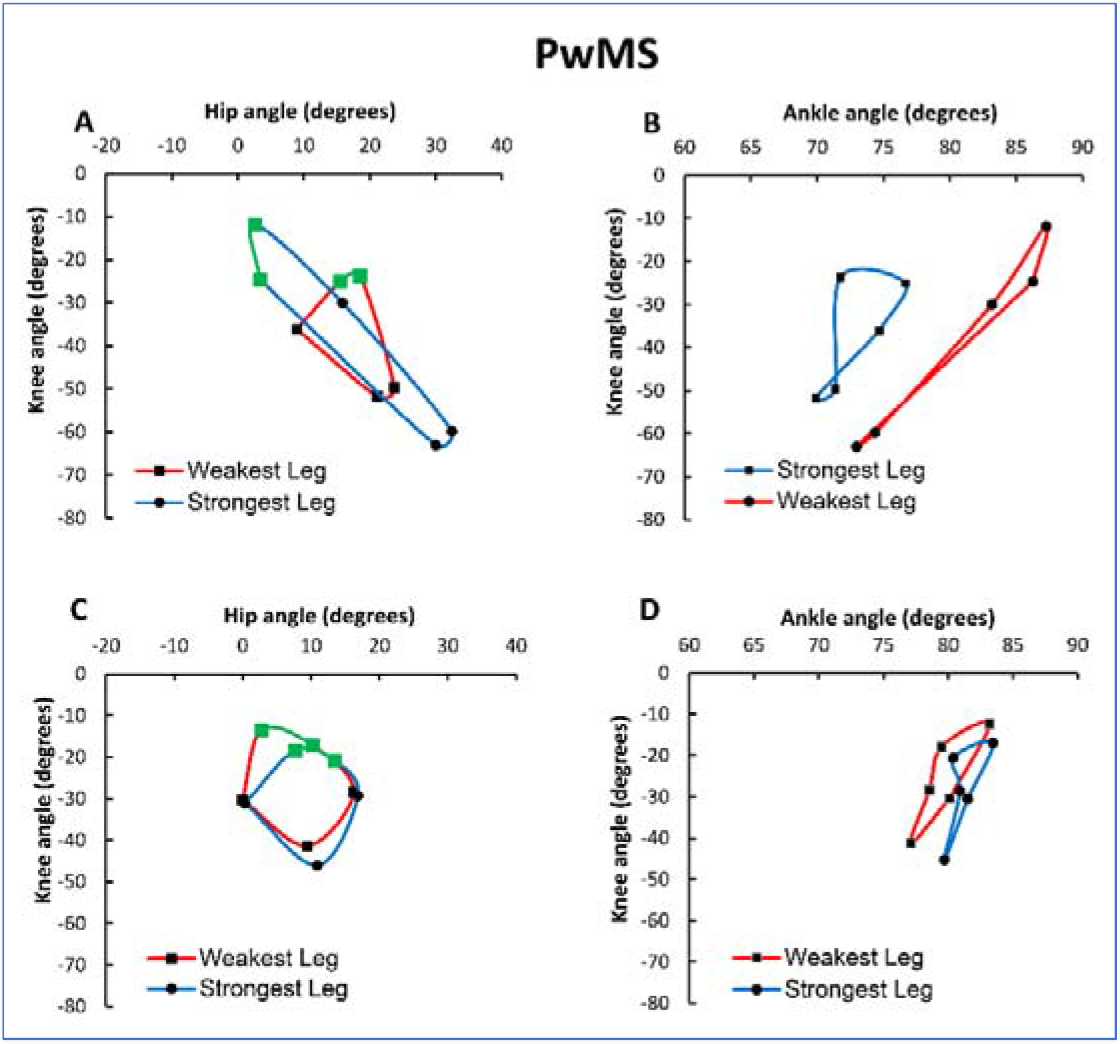
Angle-angle plots of PwMS. A and B are representative examples of one participant; C and D are the mean values of the PwMS group. The green lines indicate the phases between foot strike (FS) and maximal knee flexion during the stance phase (MKF-C).

### EMG Activity

The sEMG_RMS_ activities in the leg muscles did not show significant differences among the ten contact phases in either group (P>0.05); however, in PwMS, the bar errors were larger than those of the controls (Fig 6). Activation of the vastus lateralis was significantly higher in PwMS than in HCs, even when they walked at higher speeds (PwMS vs. HC-1, P<0.001, ES=1.89; PwMS vs. HC-3, P<0.01, ES=1.80; Fig 6A). Similarly, biceps femoris muscle activation was higher in PwMS than in HC-1 (P<0.05, ES=1.30; Fig 6B). In contrast, PwMS showed less activation in the tibialis anterior (PwMS vs. HC-1, ES=1.76; P<0.01; Fig 6C) and lateral gastrocnemius muscles (PwMS vs. HC-1, P<0.01, ES=1.70; Fig 6D). By increasing the speed from 1 km/h to 3 km/h, HCs increased their activation in the vastus lateralis (HC-3 vs. HC-1, P<0.05, ES=1.91; Fig 6A), biceps femoris (HC-3 vs. HC-1, P<0.001, ES=1.87; Fig 6B) and lateral gastrocnemius muscles (HC-3 vs. HC-1, P<0.05, ES=1.58; Fig 6D). In contrast, the tibialis anterior did not increase its activation at higher speeds (HC-3 vs. HC-1, P>0.05; Fig 6C).

## Discussion

The main result of the present study is that the area and perimeter of PwMS were lower than those of HCs in the knee-angle/ankle-angle diagrams, whereas the differences in the hip-angle/knee angle diagrams did not reach statistical significance; therefore, our hypothesis was partially confirmed. Moreover, EMG activity revealed different leg muscle activation between the two groups during the stance phase.

Overall, these findings suggest that the angle-angle diagram, when determined between the knee and the ankle joints, is a sensitive approach to detect gait alterations in ROM and coordination in PwMS with respect to controls while walking on a treadmill at the same speed.

We are currently not aware of studies comparing the areas and perimeters of angle-angle diagrams determined at the level of hip, knee and ankle joints in PwMS and controls. Interestingly, Filli et al. (18) reported alterations in intralimb joint coordination and ROM over a period of one year by inspecting the shape and size of hip-knee and knee-angle plots in PwMS and controls; however, the analysis was qualitative without quantifying the areas and perimeters of the loops.

In the present study, quantitative analysis was possible through graphical determination of areas and perimeters in the two groups when walking at 1 km/h (PwMS and HCs) and 3 km/h (HCs); however, differences between the two groups at the same speed were computed only at 1 km/h, as the PwMS were not able to walk at speeds higher than 1 km/h. In the latter conditions, PwMS revealed a significantly reduced area (large effect size) and nearly significant perimeters compared to the HCs in the knee-angle/ankle-angle diagrams; however, they were able to maintain the goal speed of 1 km/h by increasing gait frequency.

PwMS showed high variability within subjects in the area and perimeter in the hip-knee and knee-ankle diagrams between the strongest and weakest legs during a gait cycle (CV=84-22.75%, Table 1). Additionally, the variability between subjects was high in the weakest (CV=69.10-37.00%, Table 1) and strongest legs (CV=72.16-27.77, Table 1). In a representative example of PwMS, the differences in area and perimeter between the weakest and strongest legs highlight a typical pattern (Figs 3A, 3B); specifically, the reduced conjoint range of motion (hip-knee) of the weakest leg is offset by the strongest leg, and the graph is lengthened along the positive X-axis values, indicating only forward hip rotation with large knee flexion. This pattern determined a higher perimeter, indicating that the two joints are uncoordinated during a gait cycle. A similar pattern is described by the knee-ankle diagram in which the very limited range of motion of the weakest leg is offset by the contralateral strongest leg (Fig 3B). Together, both diagrams are useful to describe the kinematic characteristics of PwMS when walking. However, as the variability between and within subjects is very high, the individual gait patterns are smoothed when averaging the values of several PwMS (Figs 3C, 3D).

Concerning the HCs, the area and perimeter were more regular between the dominant (right) and nondominant legs (left) (Figs 4A, 4B); the CVs within subjects were 31% for the area knee angle and 11.24% for the perimeter (Table 1). In contrast, the variability between subjects in HC-1 was similar to that of PwMS in terms of the area and perimeter of both diagrams (hip-knee and knee-angle) (Table 1). Therefore, when the kinematic data of HCs were averaged, the individual characteristics were less appreciable in the graphs (Figure 4C, 4D).

**Fig 4.**
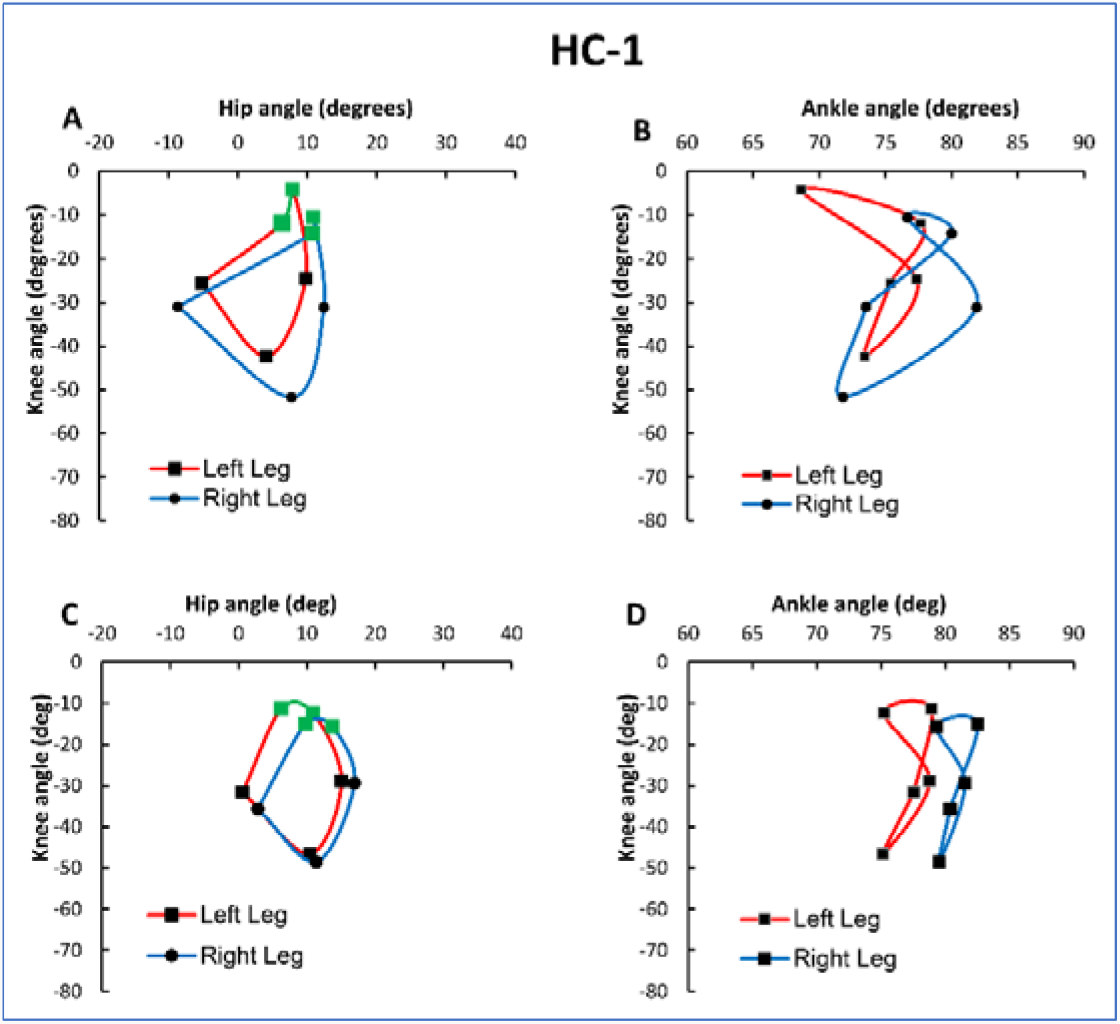
Angle-angle plots of HC-1. A and B are representative examples of one participant; C and D are the mean values of the HC group. HC-1 (speed= 1 km/h). The green lines indicate the phases between foot strike (FS) and maximal knee flexion during the stance phase (MKF-C).

The high variability between HCs can be explained by the walking speed, as 1 km/h is too low and unusual for HCs; in other words, it represents a constraint. This condition reduced the step length and the foot contact near the projection of the centre of mass on the ground; consequently, knee flexion (cushioning effect) was absent, and the limb appeared to be a rigid lever in the hip-knee angle diagram (Figure 4C). In fact, in the phase from FS to MKF-C, the knee tended to extend rather than flex, similar to PwMS (Figs 3C, 3D). The low walking speed is also reflected in the knee-ankle diagram, highlighting a reduced area and perimeter (Figs 2A, 2B).

In contrast, when HCs walked at 3 km/h (i.e., the usual walking speed in healthy subjects) the knee flexed, and the hip extended between FS and MKF-C. Additionally, the shape and dimension of hip-knee and knee-ankle diagrams in the representative example (Figs 5A, 5B) were similar to those of the averaged values (Figs 5C, 5D). Consequently, the variability within and between subjects decreased by increasing the speed walking, particularly the CV within subjects (ranging from 6 to 14%); however, the CV of areas (hip-knee and knee-ankle diagrams) between subjects tended to remain high (ranging from □30 to 40%) (Table 1). The variability of the areas (hip-knee and knee-ankle) between the subjects was likely determined by the individual combinations selected by the subjects between step length and frequency.

**Fig 5.**
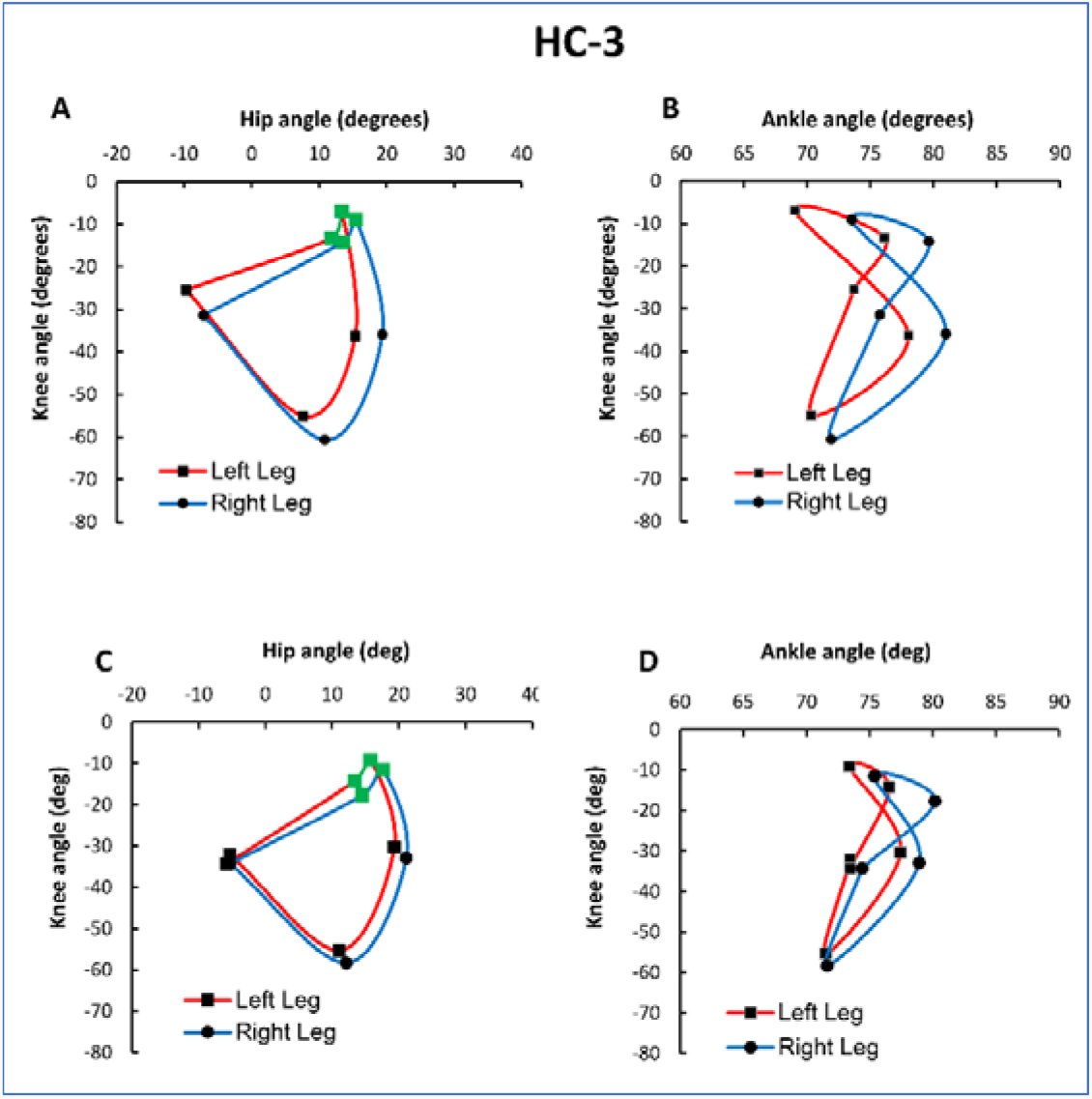
Angle-angle plots of HC-3 (speed=3 km/h). A and B are representative examples of one participant; C and D are the mean values of all participants in the HC group. The green lines indicate the phases between foot strike (FS and maximal knee flexion during the stance phase (MKF-C).

**Fig 6.**
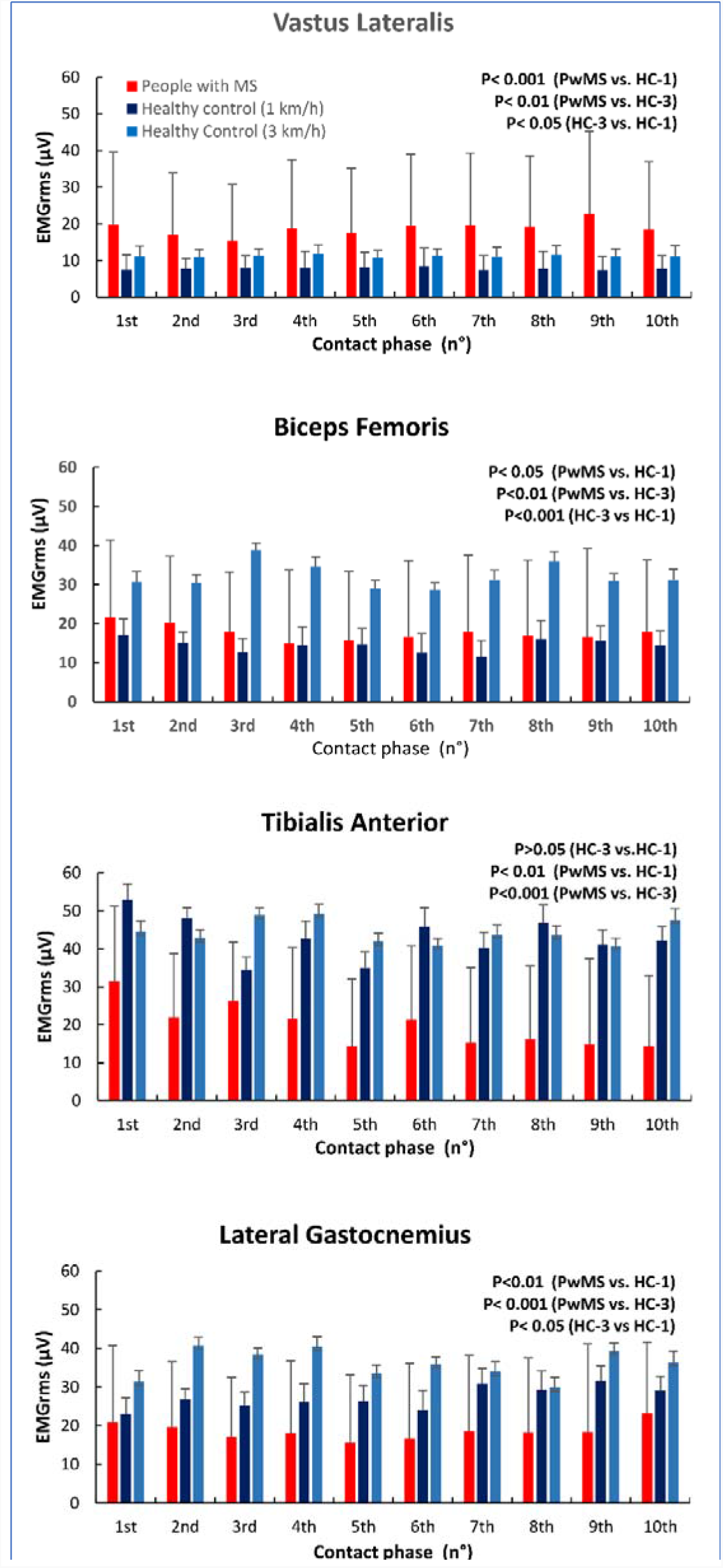
Average normalized sEMGrms (SD) values of leg muscles (vastus lateralis, biceps femoris, tibialis anterior and lateral gastrocnemius) recorded for the weakest lower limb in PwMS (1 km/h) and for the dominant limb in HCs (1 km/h and 3 km/h). The significant differences between the two groups are reported in the text boxes within the graphs.

These results are consistent with the activation differences in proximal (VL and BF) and distal (TA and LG) leg muscles between PwMS and HC. In the present study, the high activation of the VL and BF indicates a stiff knee during the stance phase (appreciable in the hip-knee diagram, green line), which causes PwMS to compensate for the deficit with the ankle muscles (TA and LG). The reduced muscle activation of the TA and LG is responsible for nonfunctional limb distal control, which alters ankle positioning in the initial phase of the stride and the subsequent propulsion, thereby reducing the contribution to the forwards momentum of the body (19). Therefore, altered muscle activation could be explained by a different recruitment pattern of motor units in proximal vs. distal muscles (20) and represents a compensatory strategy that could increase when the balance is impaired in PwMS (21).

The potential points of concern in the present study include the following. First, the sample size was relatively small (N=20), and post hoc analysis demonstrated that the computed statistical power was equal to 0.16-0.19 to detect a small effect size (□0.10) and a significant difference (p<0.05) between PwMS and HCs when perimeters and areas were determined in the hip-knee angle. The achieved power of 0.16-0.19 corresponds to a likelihood of only 16-19% for revealing a real difference with a sample size of N=200; therefore, a larger sample size would have been unlikely to lead to a different result, indicating that the more sensitive parameter to detect gait alteration in PwMS is the knee-ankle diagram.

Second, EMG activity was recorded in the muscles of only one leg (the weakest in PwMS and the dominant in HCs). Regarding the latter limitation, we emphasize that recording the EMG activity in both leg muscles would make it possible to better describe the pattern of activation and the interactions between the weakest and strongest leg muscles during the gait cycles.

To summarize, the results of the present study confirm the feasibility of angle-angle diagrams to provide individual quantitative and qualitative information regarding ROM and coordination while PwMS are walking. Future investigations should confirm this preliminary study by exploring the sensitivity of the angle-angle diagram to describe and quantify gait modifications in longitudinal protocols. Therefore, it is reasonable to suggest that the information provided by the angle-angle diagram could be used to detect individual gait alterations and assess the effectiveness of exercise and/or pharmacological interventions in PwMS.

## Data Availability

All data produced in the present study are available upon reasonable request to the authors

## Institutional Review Board Statement

The study was conducted according to the guidelines of the Declaration of Helsinki and approved by the Internal Review Board of the University (Protocol code n°13/2021).

## Informed Consent Statement

Informed consent was obtained from all subjects involved in the study.

## Data Availability Statement

The data presented in this study are available on request from the corresponding authors.

## Acknowledgments

The authors would like to thank Federico Di Domenico for his support in the areas and perimeters computation by using the software Auto-CAD.

